# Hospital admissions for acute respiratory tract infections among infants from Nunavut and the burden of respiratory syncytial virus: a 10-year review in regional and tertiary hospitals

**DOI:** 10.1101/2024.02.21.24303174

**Authors:** Mai-Lei Woo Kinshella, Jean Allen, Jasmine Pawa, Jesse Papenburg, Radha Jetty, Rachel Dwilow, Joanne Embree, Joan Robinson, Laura Arbour, Manish Sadarangani, Ye Shen, Jeffrey N. Bone, Celia Walker, Iryna Kayda, Holden Sheffield, Darcy Scott, Amber Miners, David M. Goldfarb

**Affiliations:** BC Children’s Hospital Research Institute, Vancouver, British Columbia, Canada; Nunavut Tunngavik Incorporated, Iqaluit, Nunavut, Canada; Department of Health, Government of Nunavut, Iqaluit, Nunavut, Canada; Division of Clinical Sciences, NOSM University, Ontario, Canada; Dalla Lana School of Public Health, University of Toronto, Ontario, Canada; Division of Pediatric Infectious Diseases, Department of Pediatrics, Montreal Children’s Hospital, Montreal, Canada; Division of Microbiology, Department of Clinical Laboratory Medicine, Optilab Montreal, McGill University Health Centre, Montreal, Canada; Department of Pediatrics, University of Ottawa, Ottawa, Canada; Department of Pediatrics, University of Manitoba, Winnipeg, Canada; Department of Pediatrics, University of Alberta, Edmonton, Canada; Department of Medical Genetics, Faculty of Medicine, University of British Columbia; Department of Pediatrics, Faculty of Medicine, University of British Columbia; Department of Pathology and Laboratory Medicine, University of British Columbia, Vancouver, Canada; Qikiqtani General Hospital, Iqaluit, Nunavut, Canada; Stanton Territorial Hospital, Yellowknife, Northwest Territories, Canada; Division of Medical Microbiology, BC Children’s Hospital & BC Women’s Hospital & Health Centre, Vancouver, British Columbia, Canada

## Abstract

**Background:** Nunavut is a northern Canadian territory in Inuit Nunangat (Inuit homeland in Canada). Approximately 85% of the population identifies as Inuit. A high proportion of infants in Nunavut are admitted to hospital with acute respiratory tract infection (ARI) but previous studies have been limited in regional and/or short duration of coverage. This study aimed to estimate the incidence rate, microbiology and outcomes of ARI hospitalizations in Nunavut infants.

**Methods:** We conducted chart reviews with a retrospective cohort of infants aged <1 year from Nunavut at six Canadian hospitals, including two regional and four tertiary pediatric hospitals January 1, 2010, to June 30, 2020. Descriptive statistics and multivariable logistic regression were performed.

**Results:** We identified 1189 ARI admissions of infants during the study period, with an incidence rate of 133.9 per 1000 infants per year (95% confidence interval (CI): 126.8, 141.3). Of these admissions, 56.0% (n=666) were to regional hospitals alone, 72.3% (n=860) involved hospitalization outside of Nunavut, 15.6% (n=185) were admitted into intensive care, and 9.2% (n=109) underwent mechanical ventilation. Of the 730 admissions with a pathogen identified, 45.8% had respiratory syncytial virus (RSV; n=334), for a yearly incidence rate of 37.8 hospitalizations per 1000 infants (95% CI: 33.9, 42.1). Among RSV hospitalizations, 41.1% (n=138) were infants 0-2 months of age and 32.1% (n=108) were > 6months.

**Interpretation:** Understanding the high burden of ARI among Nunavut infants can inform health policy and serve as a baseline for assessing the impact of any new interventions targeting infant ARIs.

## Introduction

Nunavut, the largest region in Inuit Nunangat, is a northern Canadian territory where approximately 85% of the population identifies as Inuit. Although there have been improvements in the management of children requiring admission to hospital with acute respiratory tract infection (ARI) (1), a high proportion of infants in the territory have historically been hospitalized due to these infections (2–4). The 2007-2008 Nunavut Inuit Child Health Survey reported 33% of children younger than two years experienced a severe lower respiratory tract infection, of which 81% resulted in hospitalizations (5). Respiratory Syncytial Virus (RSV) is the most common cause of severe ARI in infants worldwide (6) and may contribute 30-41% of ARI hospitalizations among infants from Nunavut (7,8).

Due to the remoteness of communities (all are fly-in, without road access) and complex referral pathways in Nunavut, infants admitted to hospital with RSV and other respiratory pathogens often require transport out of territory and have a high rate of intensive care admission (5). A review of pediatric urgent air transfers for children across Northern Canada found 54% of cases originated from Nunavut, where an estimated 40.7 per 1000 infants are air transported to tertiary pediatric hospitals for severe ARIs (8). Transport outside of their home community is disruptive to the whole family, which is magnified with transport out of territory.

While previous studies in Nunavut highlighted infant RSV as a concern of high priority, understanding the burden of ARI hospitalizations has been limited by coverage only in certain regions (4,9), a single season (7,9), or only those air transferred to tertiary pediatric hospitals (8). Particularly given the potential of new interventions to prevent severe RSV disease in infancy (10), the objective of our study is to comprehensively estimate the burden of ARI hospitalizations in infants from Nunavut over 10 years (2010–2020), including determining the rate, microbiology and outcomes of ARI hospitalizations.

## Methods

### Study design and population

This study is part of the larger Burden Ethnographic Modeling Evaluation Qaujilisaaqtuq (BEMEQ) RSV initiative to estimate the burden of RSV, perceptions and potential impact of preventive interventions. We conducted a retrospective chart review at six hospitals serving Nunavut’s population, including two regional hospitals (Qikiqtani General Hospital, QGH; Stanton Territorial Hospital, STH) and four tertiary care hospitals (Stollery Children’s Hospital, SCH; Winnipeg Children’s Hospital, WCH; Children’s Hospital of Eastern Canada, CHEO; Montreal Children’s Hospital, MCH). Nunavut has three regions: for the Kitikmeot region, out-of-territory referrals tend to flow to STH and SCH; for the Kivalliq region, to WCH; and for the Qikiqtaaluk/Qikiqtani region, to CHEO and MCH. This study did not capture outpatient visits to health centres.

Our study included all children <12 months of age who were residents of Nunavut with suspected or proven community-onset ARI as the reason for hospitalization admitted between January 1, 2010 and the end of the 2019-2020 RSV season (June 30, 2020). ARIs were defined as a new illness (<14 days duration) with respiratory symptoms suspected to be caused by infection. Hospitalizations were defined as admissions of at least 24 hours at the hospital centre. Primary care visits, non-ARI admissions and hospital visits less than 24 hours (i.e. emergency room visits) were excluded.

### Data collection and analysis

Hospital charts were identified using respiratory ICD-10 codes (**Supplementary file 1**) extracted from hospitalization databases at tertiary hospitals. At regional hospitals, all admissions of infants from Nunavut apart from birth hospitalizations were reviewed. Charts were reviewed by trained members of the study team in each hospital. Relevant data was extracted using a modified version of the previously utilized Urgent Air Transfer data collection form (8). When more than one pathogen was identified, the site investigator adjudicated the likely primary pathogen based on timing, site of detection and the clinical picture. Data from tertiary sites from 2010-2014 were already collected during the Urgent Air Transfer study and the current BEMEQ study added to the dataset by collecting 2015-2020 data. Regional hospitals were not included in the Urgent Air Transfer study so data were collected for 2010-2020.

The data was stored in REDCap (Research Electronic Data Capture), managed at the BC Children’s Hospital Research Institute. Descriptive statistics were conducted to present the demographics, underlying comorbidities, clinical care during admissions, and the identified microbial etiologies. Nunavut Bureau of Statistics population data were used to calculate the incidence of infant ARI and RSV hospitalization rates, based on an annual average of admissions by population over 2010 to 2019 (11). We did not calculate the incidence for 2020 due to a truncated year ending after the 2019-2020 RSV season in June and the potential influence of the COVID-19 pandemic. Logistic regression analysis was used to investigate associations between demographic characteristics and clinical outcomes firstly, among admitted infants with laboratory confirmed RSV versus other respiratory viral pathogens, and secondly, among admitted infants with RSV only versus RSV with documented coinfections (other respiratory viruses). We present result as odds ratios (ORs) and 95% confidence intervals adjusted for *a priori* identified confounders: age, time period 2010-2014/ 2015-2020, preterm birth (yes/no), and region of residence. Analyses were conducted using R statistical software.

### Ethical considerations and stakeholder engagement

The study received ethics approval from the University of British Columbia Children’s and Women’s Research Ethics Board (H19-03793), the University of Alberta Health Research Ethics Board (Pro00103234), University of Manitoba Health Research Ethics Board (H2021:259), the Children’s Hospital of Eastern Ontario Research Ethics Board (21/18X), and the Pediatric Panel of the Research Ethics Board of the Research Institute of the McGill University Health Centre (2021–6256). The study also obtained Scientific Research Licences from the Northwest Territories (16813) and Nunavut (01 011 23R-M). Exact numbers in data categories with less than five patients were not reported to protect confidentiality. Our project involved the collaboration and engagement of partners from Nunavut Tunngavik Incorporated (NTI), a representative organization for Inuit in Nunavut, and Nunavut Department of Health, including public health stakeholders and clinicians. Partners were involved throughout the study in project planning, project updates and interpretation of results, to ensure alignment with Nunavut priorities and needs.

## Results

During the study period, there were 1189 ARI admissions among infants from Nunavut (**Figure 1**), including 666 (56.0%) to regional hospitals alone, 494 (41.5%) to tertiary hospitals alone, and 29 (2.4%) first admitted at a regional hospital before being transferred to a tertiary hospital.

**Figure 1.**
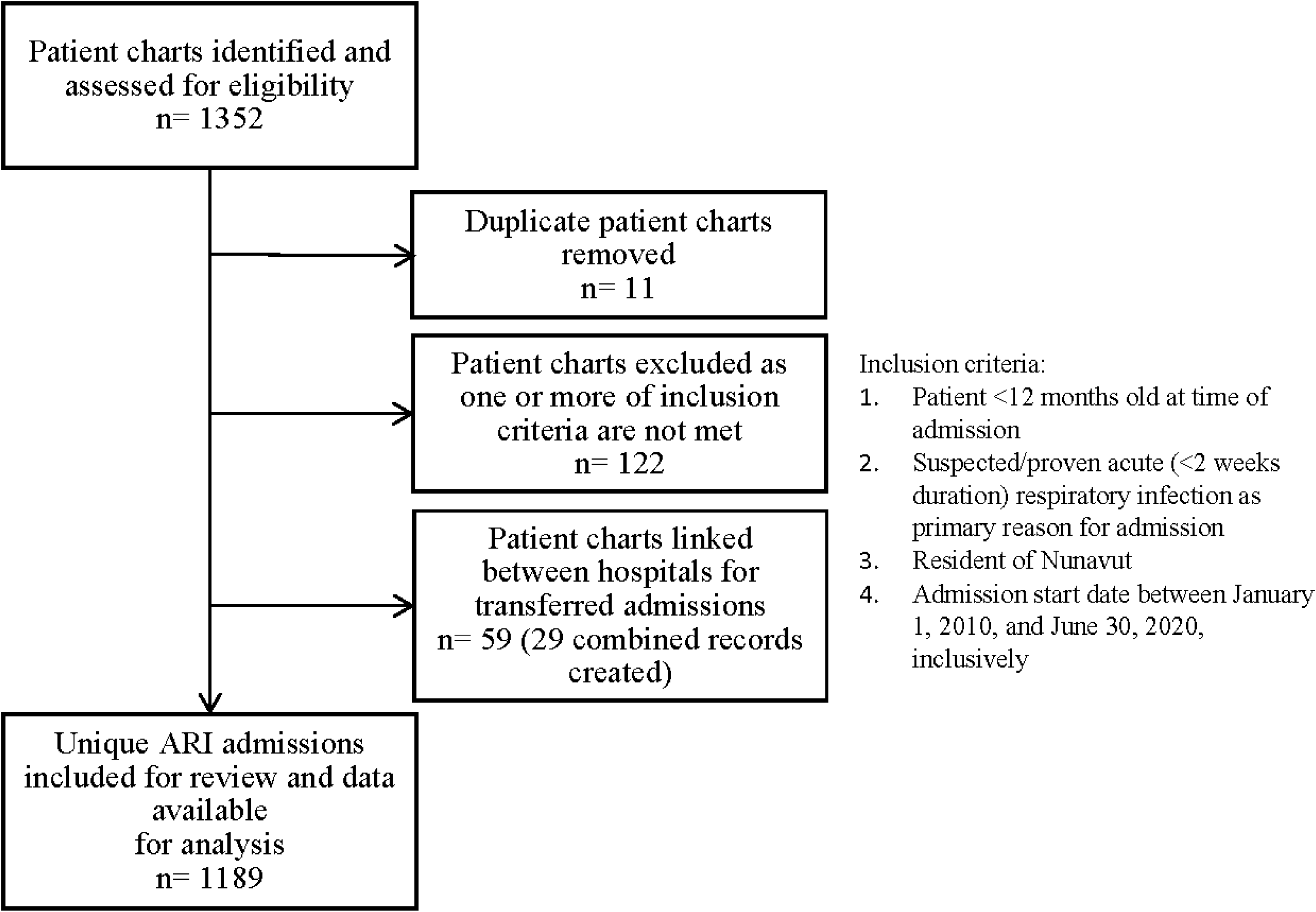
Flow diagram of patient charts.

**Figure 2.**
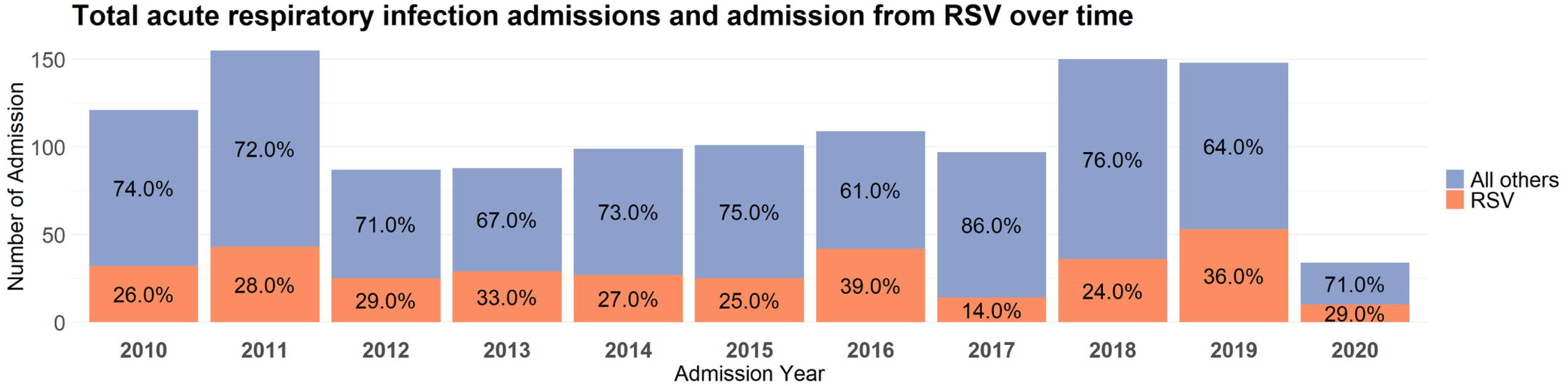
Total acute respiratory infection admissions and admissions due to RSV over time.

Three-hundred and twenty-nine infants’ (27.7%) were admitted to QGH, while the other 860 (72.3%) admissions involved hospitalization outside of Nunavut. Of the 1189 admissions, 534 (44.9%) were from the Qikiqtaaluk/Qikiqtani region, 379 (31.9%) from the Kitikmeot region, and 276 (23.2%) from the Kivalliq region.

**Table 1** summarizes the demographics of patients by hospital and overall. There were 804 (67.6%) infants less than six months of age and 385 (32.4%) six months or older. Among those admitted only to regional hospitals, 36.3% (242 of 666) were six months or older, compared to 27.3% (143 of 523) of those admitted to or transferred to the four tertiary hospitals. The median age of admission was three months, with an interquartile range of two to seven months. Almost all (968 of 972; 99.6%) infants with ethnicity recorded in their hospital charts were Inuit, 679 (57.1%) were male, and 264 (22.7%) were born premature. The median gestational age for premature infants was 35 weeks (late preterm). Underlying chronic medical conditions were reported in 205 (17.2%) cases, including 53 (4.5%) with significant cardiac and/or respiratory condition.

**Table 1.**
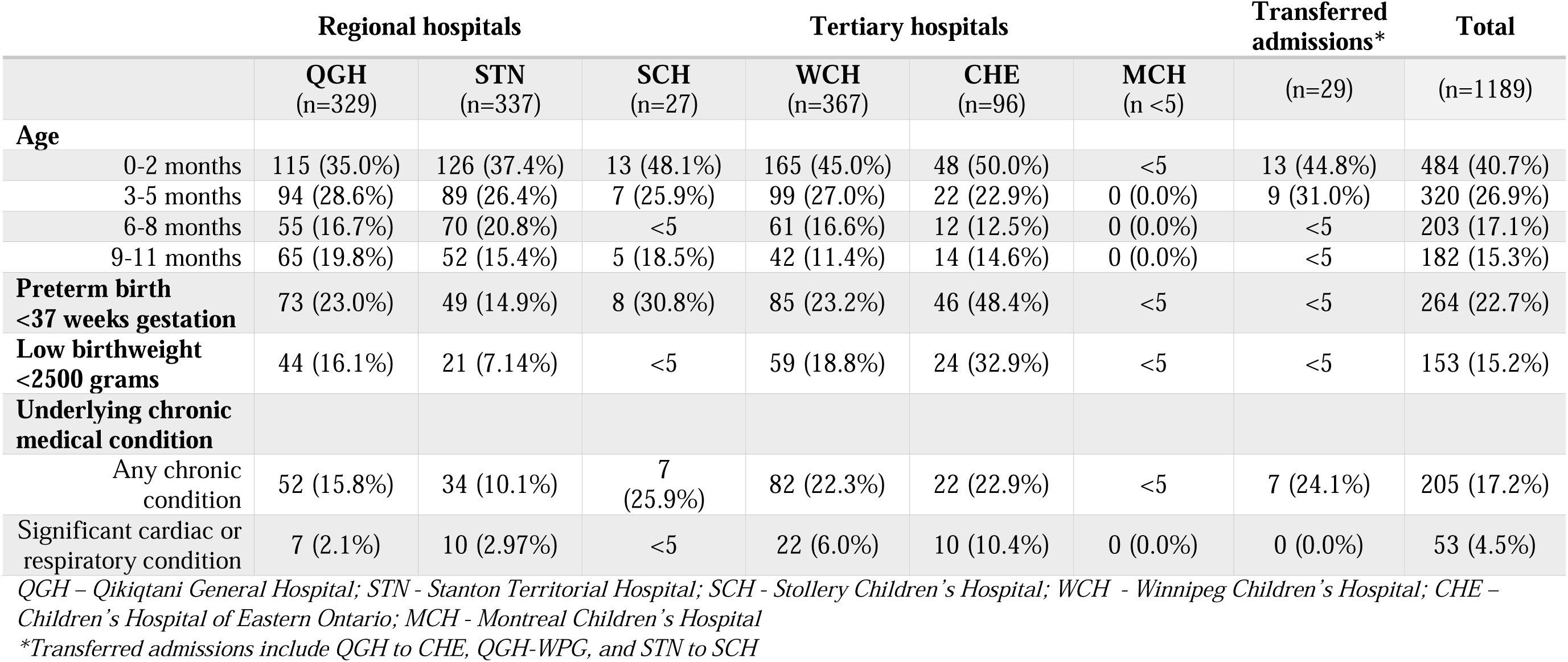
Summary of demographics for Nunavut acute respiratory infection infant admissions between 2010 and 2020.

**Table 2** summarizes admissions and outcomes by hospital. Of the 1189 admissions, 185 (15.6%) were admitted into intensive care and 109 (9.2%) were intubated and underwent mechanical ventilation. The median length of stay was five days, with lengthier stays reported at tertiary hospitals, and 381 (32.0%) were hospitalized for over a week. Pneumonia and bronchiolitis were the primary diagnosis in 1035 (87.0%) admissions. Almost all infants (1167 of 1189; 98.1%) were discharged home, though 96 (8.1%) were readmitted at a hospital within 30 days of discharge. Two infant deaths at tertiary hospitals were reported.

**Table 2.** Hospital admission and outcomes.

|  | Regional hospitals |  |  | Tertiary hospitals |  |  | Transferred admissions* | Total |
| --- | --- | --- | --- | --- | --- | --- | --- | --- |
|  | QGH<br>(n=329) | STN<br>(n=337) | SCH<br>(n=27) | WCH<br>(n=367) | CHE<br>(n=96) | MCH<br>(n <5) | (n=29) | (n=1189) |
| Admission into intensive care | 0 (0.0%) | 0 (0.0%) | 21 (77.8%) | 78 (21.3%) | 65 (67.7%) | <5 | 17 (58.6%) | 185 (15.6%) |
| Highest respiratory support |  |  |  |  |  |  |  |  |
| Oxygen only | 153 (46.5%) | 226 (67.1%) | 10 (37.0%) | 206 (56.1%) | 18 (18.8%) | 0 (0.0%) | 13 (44.8%) | 626 (52.6%) |
| CPAP/BiPAP | 0 (0.0%) | 0 (0.0%) | 0 (0.0%) | 27 (7.4%) | 22 (22.9%) | <5 | <5 | 52 (4.4%) |
| Mechanical ventilation / HFOV | 0 (0.0%) | <5 | 16 (59.3%) | 39 (10.6%) | 37 (38.5%) | <5 | 11 (37.9%) | 109 (9.2%) |
| Antimicrobial therapy | 212 (64.4%) | 237 (70.3%) | 25 (92.6%) | 230 (62.7%) | 80 (83.3%) | <5 | 26 (89.6%) | 814 (68.5%) |
| Median length of stay [IQR] | 4.0<br>[3.0; 6.0] | 5.0<br>[4.0; 8.0] | 12.0<br>[9.0; 15.0] | 5.0<br>[3.0; 9.0] | 12.0<br>[7.0; 20.0] | 16.0<br>[13.0;18.5] | 14.0<br>[11.0;22.0] | 5.0<br>[3.0;9.0] |
| Length of stay >7 days | 47 (14.3%) | 85 (25.2%) | 22 (81.5%) | 125 (34.1%) | 70 (72.9%) | <5 | 28 (96.6%) | 381 (32.0%) |
| Primary discharge diagnosis |  |  |  |  |  |  |  |  |
| Pneumonia | 61 (18.5%) | 87 (25.8%) | <5 | 86 (23.4%) | 24 (25.0%) | 0 (0.0%) | 8 (27.6%) | 269 (22.6%) |
| Bronchiolitis | 187 (56.8%) | 218 (64.7%) | 18 (66.7%) | 273 (74.4%) | 49 (51.0%) | <5 | 17 (58.6%) | 766 (64.4%) |
| Hospital readmission within 30 days | 31 (9.4%) | 20 (5.9%) | 0 (0.0%) | 31 (8.5%) | 10 (10.4%) | 0 (0.0%) | <5 | 96 (8.1%) |

QGH – Qikiqtani General Hospital; STN - Stanton Territorial Hospital; SCH - Stollery Children's Hospital; WCH - Winnipeg Children's Hospital; CHE – Children's Hospital of Eastern Ontario; MCH - Montreal Children's Hospital; CPAP – continuous positive airway pressure; BiPAP - bilevel positive airway pressure; IQR – interquartile range; HFOV - high-frequency oscillatory ventilation
\*Transferred admissions include QGH to CHE, QGH-WPG, and STN to SCH

**Table 3** summarizes the microbiology of ARI admissions. Of the 730 (61.4%) admissions with laboratory confirmation of etiology, RSV was the leading primary pathogen with 334 (45.8%) cases. There were two admissions that reported RSV as a concurrent infection to the primary pathogen. Other major pathogens reported include influenza A and B, parainfluenza, hMPV, coronavirus, adenovirus, rhinovirus, enterovirus and haemophilus influenza. There were 220 (18.5%) admissions with documented co-infections.

**Table 3.** Microbiology.

|  | Regional hospitals |  |  | Tertiary hospitals |  |  | Transferred admissions* | Total |
| --- | --- | --- | --- | --- | --- | --- | --- | --- |
|  | QGH<br>(n=329) | STN<br>(n=337) | SCH<br>(n=27) | WCH<br>(n=367) | CHE<br>(n=96) | MCH<br>(n <5) | (n=29) | (n=1189) |
| **Lab confirmation of pathogen** | 148 (45.0%) | 217 (64.4%) | 24 (88.9%) | 242 (65.9%) | 71 (74.0%) | <5 | 24 (82.8%) | 730 (61.4%) |
| **Primary pathogen identified** |  |  |  |  |  |  |  |  |
| RSV | 63 (42.6%) | 98 (45.2%) | 7 (29.2%) | 120 (49.6%) | 31 (43.7%) | <5 | 13 (44.8%) | 334 (45.8%) |
| Influenza A | 9 (6.1%) | <5 | 0 (0.0%) | 10 (4.1%) | <5 | 0 (0.0%) | 0 (0.0%) | 27 (3.7%) |
| Influenza B | <5 | <5 | 0 (0.0%) | 5 (2.1%) | 0 (0.0%) | 0 (0.0%) | 0 (0.0%) | 8 (1.1%) |
| hMPV | 13 (8.8%) | 22 (10.1%) | <5 | <5 | 5 (7.0%) | 0 (0.0%) | 0 (0.0%) | 43 (5.9%) |
| Parainfluenza | 12 (8.1%) | 22 (10.1%) | 6 (25.0%) | 14 (5.8%) | <5 | <5 | <5 | 60 (8.2%) |
| Coronavirus | 6 (4.1%) | 6 (2.8%) | 0 (0.0%) | 6 (2.5%) | <5 | 0 (0.0%) | <5 | 22 (3.0%) |
| Adenovirus | 7 (4.7%) | <5 | <5 | 18 (7.4%) | <5 | 0 (0.0%) | <5 | 34 (4.7%) |
| Rhinovirus/<br>Enterovirus | 30 (9.1%) | 59 (17.5%) | 9 (33.3%) | 61 (16.6%) | 12 (12.5%) | <5 | 6 (20.7%) | 178 (15.0%) |
| Haemophilus<br>influenzae | <5 | <5 | 0 (0.0%) | <5 | <5 | 0 (0.0%) | 0 (0.0%) | 9 (1.2%) |
| **Laboratory confirmed concurrent infection** | 47 (14.3%) | 63 (18.7%) | 10 (37.0%) | 69 (18.8%) | 21 (21.9%) | 0 (0.0%) | 10 (34.5%) | 220 (18.5%) |

QGH – Qikiqtani General Hospital; STN - Stanton Territorial Hospital; SCH - Stollery Children's Hospital; WCH - Winnipeg Children's Hospital; CHE – Children's Hospital of Eastern Ontario; MCH - Montreal Children's Hospital; RSV- Respiratory syncytial virus; hMPV - Human metapneumovirus
\*Transferred admissions include QGH to CHE, QGH-WPG, and STN to SCH

The incidence rate of infant RSV hospitalizations in Nunavut was 37.8 per 1000 infants per year (95% confidence interval [CI]: 33.9-42.1), with an overall rate of ARI hospitalizations at 133.9 per 1000 infants per year (95% CI: 126.8-141.3).

**Table 4** compares demographic characteristics and clinical outcomes by infants with laboratory confirmed RSV and other respiratory viral infections. In infants admitted for an ARI, the odds of testing RSV positive was similar for older age cohorts compared to the comparison group (0-2 month old). Compared to infants with other respiratory viral infections, RSV positive infants had lower odds of prematurity status (aOR 0.59, 95% CI: 0.41-0.86, p=0.006). Clinical outcomes were generally worse amongst RSV positive cases. For example, those with RSV had higher odds of admission into intensive care (aOR 1.65, 95% CI: 1.13-2.41, p = 0.009) and oxygen therapy (aOR 2.24, 95% CI: 1.59-3.17, p<0.001). Antimicrobial therapy was lower in those with RSV (aOR = 0.59, 95% CI: 0.42-0.82, p = 0.002). RSV positive infants had higher odds of hospital stays over a week in length (aOR = 1.40, 95% CI: 1.03-1.90, p = 0.03).

**Table 4.**
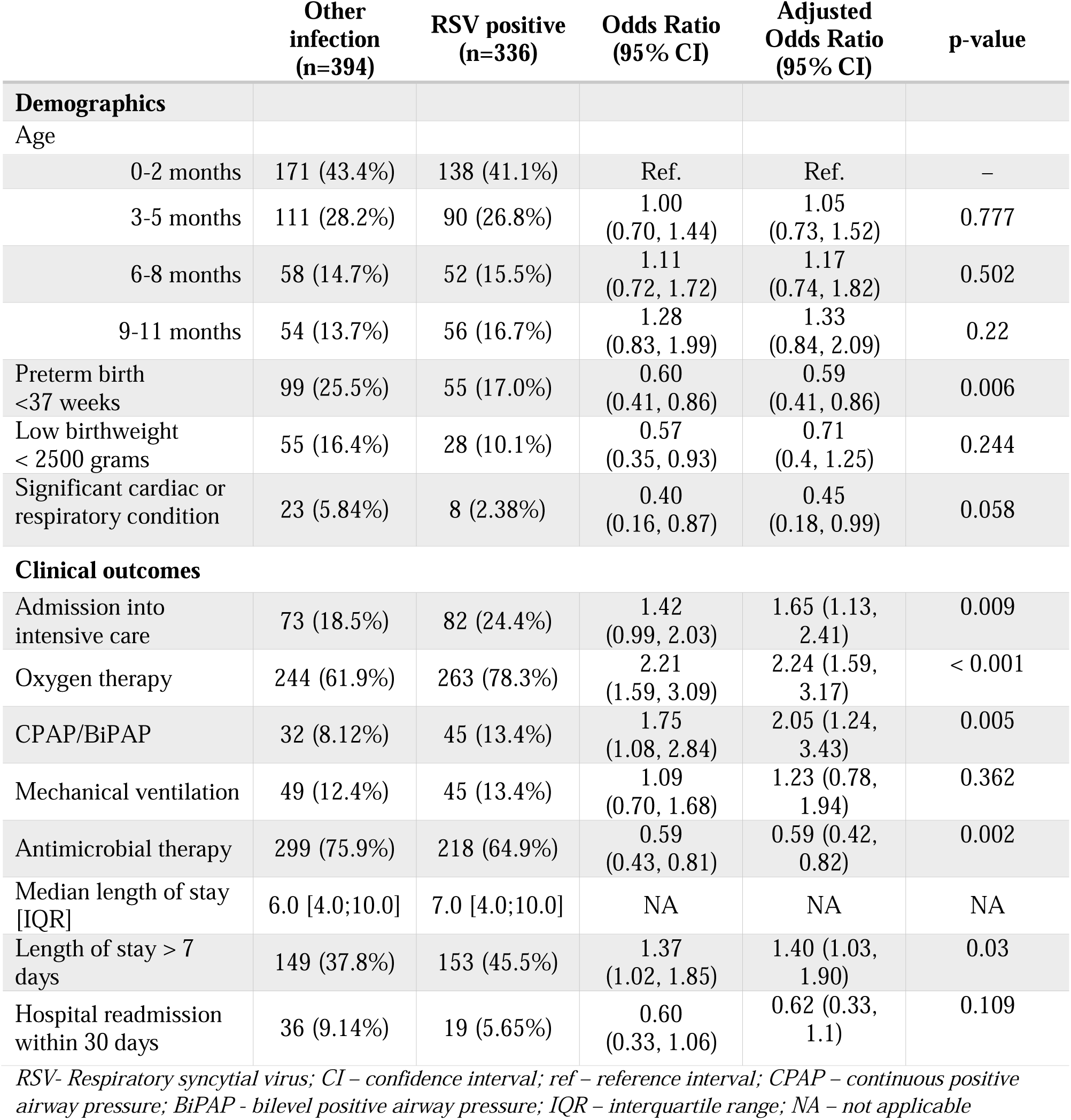
Demographic and clinical outcomes in admitted infants with laboratory confirmed RSV compared to other respiratory viral infections.

**Table 5** compares demographic characteristics and clinical outcomes for infants with RSV only versus those with RSV and concurrent infections. Concurrent infection rates were similar by infant age. Rates of low birthweight and cardiac or respiratory condition were also similar between groups. Mechanical ventilation (aOR = 2.50, 95% CI: 1.25-4.97, p = 0.009) and hospital readmission within 30 days (aOR = 3.18, 95% CI: 1.15-8.84, p =0.024) were more frequent in those with concurrent infection. Adjusted analyses also suggested higher odds of intensive care, antimicrobial therapy, oxygen therapy but all estimates had wide confidence interval compatible with both large increases and decreases.

**Table 5.** Demographic and clinical outcomes in admitted infants with laboratory confirmed RSV only compared to RSV with concurrent infection(s)

|  | **RSV only**<br>**(n=243)** | **RSV with**<br>**concurrent**<br>**infection (n=93)** | **Odds Ratio**<br>**(95% CI)** | **Adjusted**<br>**Odds Ratio**<br>**(95% CI)** | **p-value** |
| --- | --- | --- | --- | --- | --- |
| **Demographics** |  |  |  |  |  |
| Age |  |  |  |  |  |
| 0-2 months | 98 (40.3%) | 40 (43.0%) | Ref. | Ref. | – |
| 3-5 months | 69 (28.4%) | 21 (22.6%) | 0.75<br>(0.40, 1.37) | 0.63<br>(0.33, 1.2) | 0.166 |
| 6-8 months | 37 (15.2%) | 15 (16.1%) | 1.00<br>(0.48, 2.00) | 1.06<br>(0.5, 2.19) | 0.878 |
| 9-11 months | 39 (16.0%) | 17 (18.3%) | 1.07<br>(0.53, 2.10) | 1.2<br>(0.57, 2.46) | 0.626 |
| Preterm birth<br><37 weeks gestation | 44 (18.8%) | 11 (12.2%) | 0.61<br>(0.28, 1.20) | 0.62<br>(0.29, 1.24) | 0.196 |
| Low birthweight<br>< 2500 grams | 18 (9.09%) | 10 (12.7%) | 1.46<br>(0.61, 3.28) | 2.78<br>(0.98, 8.25) | 0.056 |
| Significant cardiac or<br>respiratory condition | 6 (2.47%) | 2 (2.15%) | 0.91<br>(0.12, 4.19) | 0.85<br>(0.12, 3.92) | 0.849 |
| **Clinical outcomes** |  |  |  |  |  |
| Admission into<br>intensive care | 56 (23.0%) | 26 (28.0%) | 1.30<br>(0.75, 2.22) | 1.47 (0.82,<br>2.62) | 0.191 |
| Oxygen therapy | 186 (76.5%) | 77 (82.8%) | 1.46<br>(0.81, 2.79) | 1.72 (0.90,<br>3.44) | 0.11 |
| CPAP/BiPAP | 34 (14.0%) | 11 (11.8%) | 0.83<br>(0.38, 1.68) | 0.94 (0.42,<br>1.99) | 0.876 |
| Mechanical ventilation | 26 (10.7%) | 19 (20.4%) | 2.14 (1.10,<br>4.09) | 2.50 (1.25,<br>4.97) | 0.009 |
| Antimicrobial therapy | 156 (64.2%) | 62 (66.7%) | 1.11<br>(0.67, 1.86) | 1.11 (0.66,<br>1.91) | 0.691 |
| Median length of stay<br>[IQR] | 6.0 [4.0;10.0] | 8.0 [5.0;15.0] | NA | NA | NA |
| Length of stay > 7 days | 105 (43.2%) | 48 (51.6%) | 1.40<br>(0.87, 2.27) | 1.42 (0.86,<br>2.34) | 0.173 |
| Hospital readmission<br>within 30 days | 10 (4.1%) | 9 (9.7%) | 2.49<br>(0.95, 6.47) | 3.18 (1.15,<br>8.84) | 0.024 |

*RSV- Respiratory syncytial virus; CI – confidence interval; ref – reference interval; CPAP – continuous positive airway pressure; BiPAP - bilevel positive airway pressure; IQR – interquartile range; NA – not applicable*

## Interpretation

We found a high rate of ARI admissions among infants from Nunavut, with RSV being the leading pathogen identified, though fortunately with a relatively low rate of in hospital mortality in the ten-year period. The underlying reasons for these high rates of hospitalization are likely multifactorial and may include social determinants of health disparities, such as high levels of food insecurity (12) and crowded housing (13), potential genetic determinants (14), or challenges in geographically remote communities where thresholds for admission/transfer differ from southern centres. On average, 13.4% of Nunavut’s infant population were hospitalized for ARIs each year; of these, seven in ten were hospitalized outside of Nunavut and approximately one in six admitted to intensive care. RSV admissions had heightened intensive care needs and longer hospital stays compared to infants admitted with other viral respiratory tract infections. RSV with another respiratory pathogen had higher rates of mechanical ventilation and hospital readmission compared to infants with RSV only.

Although RSV was the predominant pathogen, our longer term study reports variability in rates from year to year but overall lower RSV admission rates than in previous one-year studies (134 vs 166 in the Qikiqtaaluk/Qikiqtani region (15) and 195 in the Kitikmeot region (7) per 1000 infants). However, we continue to report substantially higher infant RSV hospitalization rates compared to global estimates [3.8% vs 1.6-1.8% global estimates (16–18)]. A study from Alberta found high hospital resource use and parental time burden, out-of pocket costs, lost work productivity and significant parental stress associated with infant RSV hospitalizations (19), which is heightened with additional complexities of accessing care in remote Nunavut communities. High rates of hospitalizations, frequently outside of territory, and intensive care admissions places a heavy burden on both the healthcare system and families. Language barriers are also a concern with hospitalizations outside of territory.

Previous studies concentrated on infant ARI admissions to tertiary care centers (8,20). Our findings highlight the importance of studying admissions to regional hospitals where the majority of hospitalizations occurred. Additionally, we found a higher proportion of infants six months or older at regional hospitals, compared to tertiary admissions. This may explain the larger proportion of RSV admissions with older infants in our study than previously reported. The IMPACT program at 13 Canadian tertiary pediatric hospitals reported 18.5% (1249 of 6737) of infant RSV hospitalizations were infants older than five months (20) versus 32.1% (108 of 336) in our study. The age of RSV admissions is an important factor in designing RSV prevention programs.

Monoclonal antibodies specific to RSV, including palivizumab and nirsevimab (21), have been developed and palivizumab has been implemented in Nunavut. Though our study recorded higher levels of prematurity and underlying chronic conditions than in the general population, the majority of patients admitted with RSV were healthy, full-term infants not eligible for palivizumab. The recently developed longer acting monoclonal antibody, nirsevimab, is protective against RSV for both healthy late preterm and term infants (22), including findings from recent European infant-wide implementation programs (23,24). A maternal vaccine has also recently been approved by regulatory authorities (including in Canada) with efficacy against RSV hospitalizations until at least 180 days after birth (25). The substantial rate of concurrent infections among infant ARI admissions from Nunavut may temper promising efficacy rates (26). Given the complexities of managing infant ARIs in Nunavut, a comprehensive approach is needed with medical therapies alongside continued strengthening non-invasive respiratory support, diagnostic tests, and on-site care from full-time pediatricians at regional hospitals (8,27). It will be helpful to assess impacts on patients/public, providers, costs, and outcomes in the context of stretched health systems, other illnesses and social determinants of health in Nunavut. Valuing Inuit expertise alongside Western biomedical approaches and ways of knowing is essential. Inuit have lived in this homeland for thousands of years, with rich culture and customs. Ongoing work to incorporate and align health services with the wisdom of *Inuit Qaujimajatuqangit* is important for the territory and for the health and wellbeing of Inuit.

The primary strength of the study is the 10-year period of review and admissions to smaller regional hospitals, allowing a better insight into the true burden of ARI in infants in Nunavut. Study limitations include not exploring the impacts of social determinants of health, voices of impacted families, primary care visits, and health system impacts, which is recommended for future research. Furthermore, laboratory testing was not always performed and negative tests were not recorded in our dataset. It was not possible to assess the opportunity costs or potential harms of focusing on RSV as opposed to other pathogens/health concerns.

## Conclusion

Our 10-year review of ARI hospital admissions is the largest among infants from Nunavut and included both tertiary and regional hospitals. Improved understanding of the heavy burden of respiratory tract infection among infants in Nunavut can help support health policy decision-making as well as serve as a baseline for understanding the impact of any new interventions targeting ARI in infants in the region.

## Data Availability

All data produced in the present study are available upon reasonable request to the authors

## Acknowledgements

We would like acknowledge the assistance provided by Michelle Dittrick throughout the study as well as the support of health records staff at each participating site. This work was supported by the Canadian Immunization Research Network (CIRN) through a grant from the Public Health Agency of Canada and the Canadian Institutes of Health Research (CNF 151944). The funders had no role in the study design, analysis, interpretation of the results, or decision to publish.

Manish Sadarangani is supported via salary awards from the BC Children’s Hospital Foundation and Michael Smith Health Research BC.

## Notes

### Competing Interest Statement

Dr Papenburg reported grants and personal fees from Merck, personal fees from AstraZeneca, and grants from MedImmune outside the submitted work. Dr Robinson received an Honoria from the Alberta Pharmacists' Association (RxA) for a talk on RSV September 2023 and has received consulting fees from Elsevier for peer review of FUO section of Clinical Overviews in 2023, both outside the submitted work. Dr Sadarangani has been an investigator on projects funded by GlaxoSmithKline, Merck, Moderna, Pfizer and Sanofi-Pasteur. All funds have been paid to his institute, and he has not received any personal payments. The remaining authors declare no conflicts of interest.

### Author Declarations

The Children and Women Research Ethics Board of the University of British Columbia gave ethical approval for this work.

### Summary of Updates

Edited to meet journal submission guidelines; Tables 4 and 5 revised; authors list updated

