## Supplementary file 1 for "Hospital admissions for acute respiratory tract infections among infants from Nunavut and the burden of respiratory syncytial virus: a 10-year review in regional and tertiary hospitals"

### Supplementary file 1: ICD-10 Diagnostic codes relating to ARI

**A15–A19 Tuberculosis**
o A15 Respiratory tuberculosis, bacteriologically and histologically confirmed
o A16 Respiratory tuberculosis, not confirmed bacteriologically or histologically
o A19 Miliary tuberculosis

**A30–A49 Other bacterial diseases**
o A21.2 Pulmonary tularaemia
o A22.1 Pulmonary anthrax
o A31.0 Pulmonary mycobacterial infection
o A36.0 Pharyngeal diphtheria
o A36.1 Nasopharyngeal diphtheria
o A36.0 Laryngeal diphtheria
o A37 Whooping cough
o A42.0 Pulmonary actinomycosis
o A43.0 Pulmonary nocardiosis
o A49.3 Mycoplasma infection, unspecified

**A80–B34 Viral infections**
o B01.2 Varicella pneumonia
o B20.6 HIV disease resulting in Pneumocystis pneumonia
o B25.0 Cytomegaloviral pneumonitis
o B34.2 Coronavirus infection, unspecified site

**B35-B49 Mycoses**
o B37.1 Pulmonary candidiasis
o B39 Histoplasmosis
o B40 Blastomycosis
o B44.0 Invasive pulmonary aspergillosis
o B44.1 Other pulmonary aspergillosis
o B45.0 Pulmonary cryptococcosis
o B46.0 Pulmonary mucormycosis

**B50-B64 Protozoal diseases**
o B58.3 Pulmonary toxoplasmosis
o B59 Pneumocystosis
o B960 Mycoplasma pneumoniae as the cause of diseases classified to other chapters
o B974 Respiratory syncytial virus as the cause of diseases classified to other chapters
o B97.2 Coronavirus as the cause of diseases classified to other chapters
o B97.8 Other viral agents as the cause of diseases classified to other chapters

**J00–J06 Acute upper respiratory infections**
o J00 Acute nasopharyngitis (common cold)

o J01 Acute sinusitis
o J02 Acute pharyngitis
o J03 Acute tonsillitis
o J04 Acute laryngitis and tracheitis
o J05 Acute obstructive laryngitis (croup) and epiglottitis
o J06 Acute upper respiratory infections of multiple and unspecified sites

**J09–J18 Influenza and Pneumonia**
o J09 Influenza due to identified avian influenza virus
o J10 Influenza due to identified influenza virus
o J11 Influenza, virus not identified
o J120 Adenoviral pneumonia
o J13 Pneumonia due to Streptococcus pneumoniae
o J14 Pneumonia due to Haemophilus influenzae
o J15 Bacterial pneumonia, not elsewhere classified
o J16 Pneumonia due to other infectious organisms, not elsewhere classified
o J17 Pneumonia in disease classified elsewhere
o J18 Pneumonia, organism unspecified

**J20–J22 Other acute lower respiratory infections**
o J20 Acute bronchitis
o J21 Acute bronchiolitis
o J22 Unspecified acute lower respiratory infection

**J30–J39 Other diseases of upper respiratory tract**
o J36 Peritonsillar abscess
o J390 Retropharyngeal and parapharyngeal abscess

**J840–J849 Other interstitial pulmonary diseases**
o J848 Other specified interstitial pulmonary diseases
o J849 Interstitial pulmonary disease, unspecified

**J85–J86 Suppurative and necrotic conditions of lower respiratory tract**
o J85 Abscess of lung and mediastinum
o J86 Pyothorax, Empyema

**J90–J94 Other diseases of pleura**
o J90 Pleural effusion, not elsewhere classified, Pleurisy with effusion
o J91 Pleural effusion in conditions classified elsewhere

**P20–P29 Respiratory and cardiovascular disorders specific to the perinatal period**
o P23 Congenital pneumonia
o P284 Other apnoea of newborn
